# Proteome-wide Mendelian randomization implicates nephronectin as an actionable mediator of the effect of obesity on COVID-19 severity

**DOI:** 10.1101/2022.06.06.22275997

**Authors:** Satoshi Yoshiji, Guillaume Butler-Laporte, Tianyuan Lu, Julian Daniel Sunday Willett, Chen-Yang Su, Tomoko Nakanishi, David R. Morrison, Yiheng Chen, Kevin Liang, Michael Hultström, Yann Ilboudo, Zaman Afrasiabi, Shanshan Lan, Naomi Duggan, Chantal DeLuca, Mitra Vaezi, Chris Tselios, Xiaoqing Xue, Meriem Bouab, Fangyi Shi, Laetitia Laurent, Hans Markus Münter, Marc Afilalo, Jonathan Afilalo, Vincent Mooser, Nicholas J Timpson, Hugo Zeberg, Sirui Zhou, Vincenzo Forgetta, Yossi Farjoun, J. Brent Richards

## Abstract

Obesity is a major risk factor for COVID-19 severity; however, the mechanisms underlying this relationship are not fully understood. Since obesity influences the plasma proteome, we sought to identify circulating proteins mediating the effects of obesity on COVID-19 severity in humans. Here, we screened 4,907 plasma proteins to identify proteins influenced by body mass index (BMI) using Mendelian randomization (MR). This yielded 1,216 proteins, whose effect on COVID-19 severity was assessed, again using MR. We found that a standard deviation increase in nephronectin (NPNT) was associated with increased odds of critically ill COVID-19 (OR = 1.71, *P* = 1.63 × 10^−10^). The effect was driven by an NPNT splice isoform. Mediation analyses supported NPNT as a mediator. In single-cell RNA-sequencing, *NPNT* was expressed in alveolar cells and fibroblasts of the lung in individuals who died of COVID-19. Finally, decreasing body fat mass and increasing fat-free mass were found to lower NPNT levels. These findings provide actionable insights into how obesity influences COVID-19 severity.

## Introduction

Coronavirus disease (COVID-19) has claimed more than 6 million lives globally since the beginning of the pandemic^1^, and obesity increases the risk of severe COVID-19^2,3^. To date, multiple pathways have been explored as mechanisms linking obesity to COVID-19 severity, including metabolic abnormalities, systemic inflammation, and respiratory dysfunction (e.g., impaired gas exchange)^3-5^. However, the underlying mediators whereby obesity influences COVID-19 outcomes are not fully understood. One strategy to disentangle this relationship is to identify circulating proteins mediating the effects of obesity on COVID-19 outcomes. Since circulating proteins can be measured and, in some cases modulated, the identification of mediator proteins may provide insights into the pathways whereby obesity increases the risk of severe COVID-19 and offer potential targets for therapeutic interventions.

A previous large cross-sectional study showed that body mass index (BMI) is significantly associated with changes in the plasma levels of 1,576 proteins^6^, and another study supported the considerable influence of obesity on the plasma proteome^7^. However, since proteins are intricately involved in complex biological processes, observational studies may be biased by unmeasured confounding factors and reverse causation. Considering that COVID-19 has been associated with substantial changes in the levels of circulating proteins^8^, the effects of circulating proteins on COVID-19 severity are also subject to such biases.

Mendelian randomization (MR) can help to protect against such biases. MR is a method that uses genetic variants as instrumental variables to evaluate the causal effects of exposures (risk factors) on outcomes. Since genetic variants are randomly allocated at conception, they are largely independent confounders, thereby decreasing the risk of confounding. Additionally, they are not subject to reverse causation since the allocation of genetic variants always precedes the onset of diseases^9,10^.

From the beginning of the pandemic, MR has played a critical role in providing evidence of modifiable risk factors for COVID-19^2,11,12^. For example, multiple MR studies have identified potential therapeutic targets for COVID-19, including OAS1, ABO, IFNAR2, IL-6, ELF5, and FAS^12-18^. Indeed, some MR findings have been validated by randomized controlled trials, thereby demonstrating the utility of MR^19-24^.

Nevertheless, MR is based on several instrumental variable assumptions:^9,10^ (I) the genetic variants used as instrumental variables are associated with the exposure; (II) they are not associated with factors that confound the relationship between the exposure and the outcome and; (III) they influence the outcome only through exposure (also known as exclusion restriction). The most problematic of these assumptions is the last, since the violation of exclusion restriction can bias the causal estimate through directional horizontal pleiotropy; therefore, careful assessment of directional horizontal pleiotropy is required. However, with a proper selection of instrumental variables and sensitivity analyses, MR can serve as a powerful tool to help understand causal mechanisms for disease in humans^9,10^, and in the case of obesity and COVID-19, can be used to screen thousands of proteins that may help to explain this relationship.

Here, we integrated proteome-wide MR using the large-scale aptamer-based plasma protein measurements, multiple sensitivity analyses, colocalization, fine-mapping, single-cell RNA-sequencing analysis, and mediation analysis to identify plasma proteins mediating the effect of obesity on COVID-19 severity.

## Results

### Study overview and summary

The study was conducted in the following manner (**Figure 1**):

**Figure 1.**
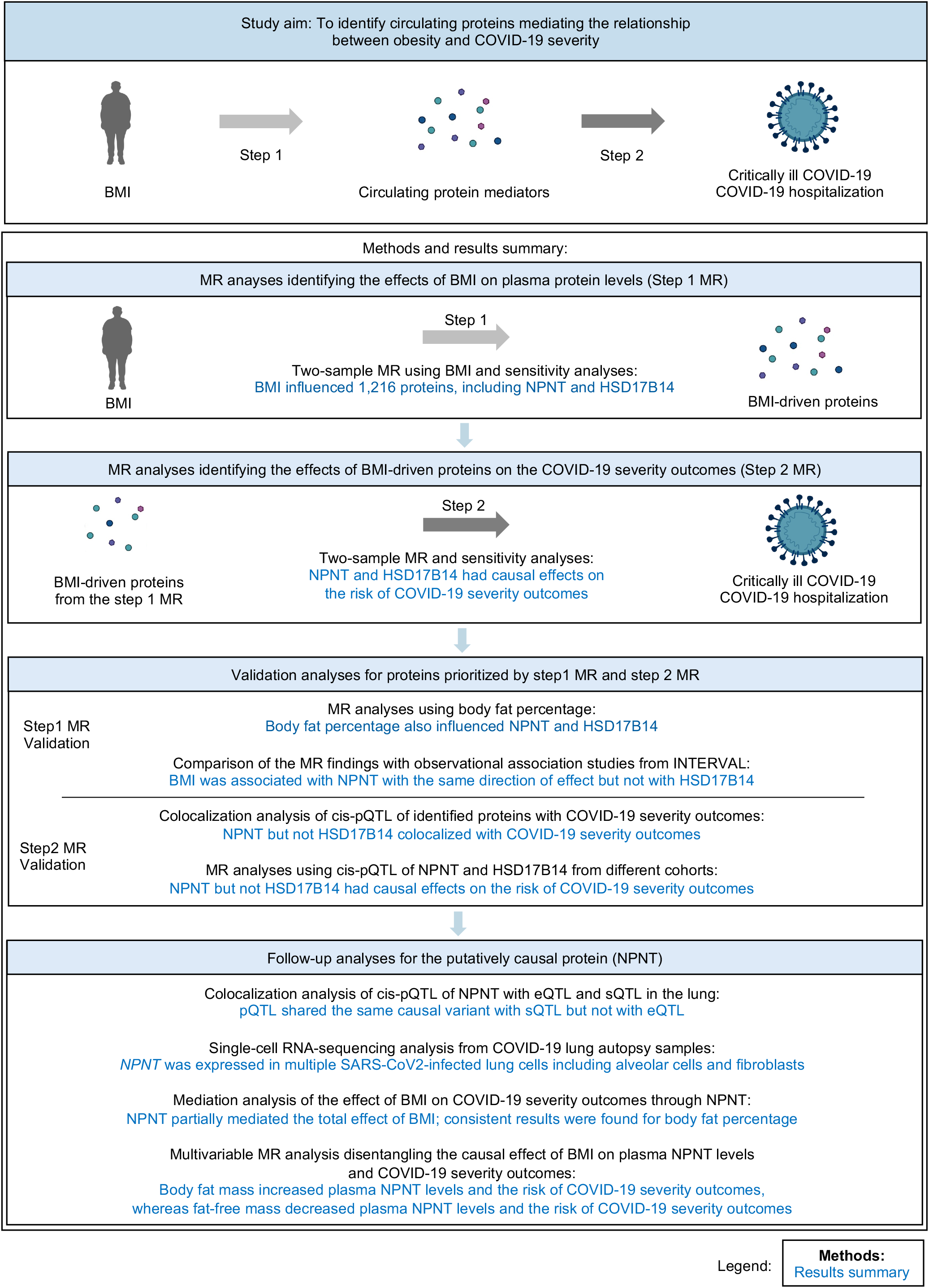
Study overview and summary. We identified circulating proteins mediating the effect of obesity on COVID-19 severity using two-step MR approach: First, we estimated the effect of BMI on 4,907 plasma proteins using MR, which yielded 1,216 BMI-driven proteins (Step 1 MR). Second, we estimated the effect of the BMI-driven proteins on COVID-19 severity outcomes, again using MR (Step 2 MR). This was followed by multiple validity assessments and follow-up analyses. MR: Mendelian randomization, BMI: body mass index, NPNT: nephronectin, HSD17B14: hydroxysteroid 17-beta dehydrogenase 14, cis-pQTL: cis-acting quantitative trait loci, e-QTL: expression quantitative trait loci, sQTL: splicing quantitative trait loci.

#### 1) Step 1 MR

First, we estimated the effect of BMI on circulating protein levels using two-sample MR on a proteome-wide scale. Two-sample MR can estimate the causal effect of the exposure on the outcome using summary statistics of genome-wide association studies (GWAS). For this, we used a GWAS of BMI in 694,649 individuals from GIANT and UK Biobank by Yengo *et al*.^25^, and plasma proteome GWAS from the deCODE study^26^ that measured plasma protein abundances using 4,907 aptamers in 35,559 individuals. Hereafter, “protein” is used when referring to an aptamer targeting a protein^26^. After two-sample MR and sensitivity analyses, 1,216 proteins were estimated to be influenced by BMI, including nephronectin (NPNT) and hydroxysteroid 17-beta dehydrogenase 14 (HSD17B14).

#### 2) Step 2 MR

Next, we performed two-sample MR to estimate the causal effects of the above-identified proteins (BMI-driven proteins) on critically ill COVID-19 and COVID-19 hospitalization outcomes (collectively referred to as COVID-19 severity outcomes). For this analysis, we used cis-acting protein quantitative loci (cis-pQTLs) from the deCODE study^26^, thereby minimizing the risk of directional horizontal pleiotropy. For COVID-19 severity outcomes, we used GWAS data from the COVID-19 Host Genetics Initiative^2^. This step 2 MR identified NPNT and HSD17B14 as putatively causal proteins for the COVID-19 severity outcomes.

#### 3) Validation analyses for NPNT and HSD17B14

We performed multiple validation analyses for step 1 and step 2 MR as follows:

##### 3.1. Step 1 MR validation

To validate whether NPNT and HSD17B14 were influenced by obesity, we performed separate MR analyses using body fat percentage (another proxy measure for obesity) as the exposure and plasma protein levels as the outcomes. The MR analysis found that both NPNT and HSD17B14 were increased by body fat percentage, consistent with the step 1 MR using BMI.

We also checked whether NPNT and HSD17B14 were observationally associated with BMI using a published observational association study from INTERVAL (*n* = 2,729) and found that NPNT, but not HSD17B14, was positively associated with BMI.

These validation analyses collectively supported the causal effect of obesity on NPNT, but not on HSD17B14.

##### 3.2. Step 2 MR validation

To ensure that the findings in step 2 MR were not biased by linkage disequilibrium (LD), which can reintroduce confounding, we used colocalization analyses to evaluate whether the cis-pQTLs of the identified proteins and COVID-19 severity outcomes shared a single causal variant. We found that NPNT, but not HSD17B14, shared a single causal variant with COVID-19 severity outcomes, indicating that the MR estimates for HSD17B14 in step 2 MR could have been biased by LD.

In addition, we repeated MR using cis-pQTLs from two independent studies (the FENLAND study^27^ and the AGES Reykjavik study^28^), which showed that NPNT, but not HSD17B14, influenced the COVID-19 severity outcomes. Given the lack of colocalization and replication for HSD17B14, we excluded HSD17B14 from further analyses.

For NPNT, we further investigated the consistency between MR findings and observational associations between NPNT and COVID-19 severity outcomes using the BQC19 cohort, which showed the same direction of effect as the MR analyses.

Collectively, these findings showed a causal effect of plasma NPNT levels on COVID-19 severity outcomes.

#### 4) Follow-up analyses for NPNT

##### 4.1. Colocalization of NPNT’s cis-pQTL with eQTL and sQTL

The observed differences in aptamer-measured NPNT levels may be predominantly explained by a particular isoform, rather than total NPNT levels. Thus, we used colocalization to evaluate whether the cis-pQTL shared the same causal variant with either its expression QTL (eQTL; genetic variants that explain the total RNA expression levels of NPNT) or its splicing QTL (sQTL; genetic variants that explain a specific isoform level of NPNT). We found that the cis-pQTL shared the same causal variant with the sQTL but not with the eQTL. These findings demonstrate that an NPNT splice isoform is likely measured by the aptamer targeting NPNT.

##### 4.2. Single-cell RNA-sequencing data of SARS-CoV-2-infected lungs

Next, to gain insights into the biological role of NPNT in SARS-CoV-2-infected lungs, we analyzed single-cell RNA-sequencing data of the lung autopsy samples from patients who died due to COVID-19. We found that NPNT is significantly expressed in in alveolar cells and fibroblasts of the lung, indicating its role in air exchange and fibrosis.

##### 4.3. Mediation analysis

Further, we performed an MR mediation analysis to quantify the extent to which the total effect of obesity on COVID-19 severity outcomes was mediated by plasma NPNT levels. We found that NPNT partially mediated the total effect of BMI, and consistent results were found for body fat percentage.

##### 4.4. Multivariable MR analyses of body fat and fat-free mass

Finally, we estimated the independent causal effect of body fat and fat-free mass on plasma NPNT levels and COVID-19 severity outcomes using multivariable MR. We found that body fat mass increased plasma NPNT levels and the risk of COVID-19 severity outcomes, whereas fat-free mass decreased plasma NPNT levels and the risk of COVID-19 severity outcomes. These findings demonstrate that decreasing body fat and increasing body fat mass (e.g., through actions such as appropriate exercise and diet) can decrease plasma NPNT levels, and thus, may reduce COVID-19 severity outcomes, thereby suggesting NPNT as an actionable target.

Each of these steps is described in more detail below.

#### 1. BMI to plasma proteins (Step 1 MR)

To estimate the causal effect of BMI on plasma protein levels on a proteome-wide scale, we performed a two-sample MR using BMI as the exposure and 4,907 plasma protein levels as the outcomes. For two-sample MR, we used an inverse variance weighted method with a random-effects model (see **Methods** and **Supplementary Table 1** for details). The F-statistic, which is a measure of the strength of the association between genetic variants and BMI, was 94.2 and thus did not indicate weak instrument bias (suspected when F-statistic < 10)^29^ (**Supplementary Tables 2**). Out of the 4,907 proteins screened, 1,304 were estimated to be influenced by BMI, using a Bonferroni-adjusted threshold of *P* < 1.0 × 10^−5^ (0.05/4907), highlighting the substantial influence of BMI on plasma protein levels (**Figure 2. Supplementary Table 3 and 4**).

**Figure 2.**
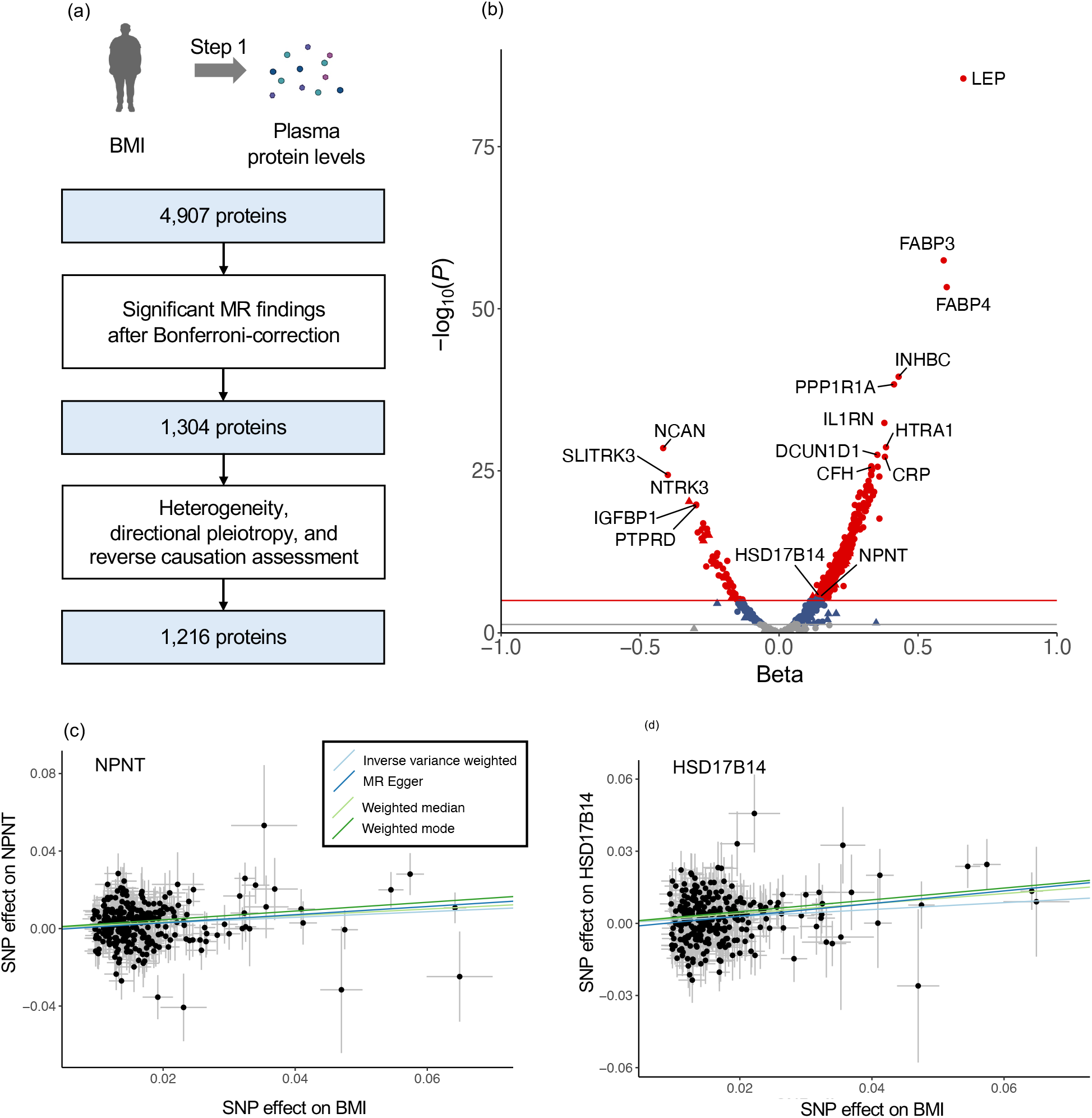
MR analyses for the effect of body mass index on plasma protein levels. (a) Flow diagram of the Step 1 MR analyses. (b) Volcano plot illustrating the effect of BMI on each plasma protein from the MR analyses using inverse variance weighted method. Red and blue horizontal lines represent *P* = 1.0 × 10^−5^ (Bonferroni correction for 4,907 proteins: 0.05/4,907) and 0.05, respectively. A proteins’ shape denotes whether the protein passed all sensitivity tests (i.e., heterogeneity, directional pleiotropy, and reverse causation assessment) (circle) or failed any of them (triangle). (c) MR scatter plot for the effect of BMI on plasma NPNT levels. (d) MR scatter plot for the effect of BMI on plasma HSD17B14 levels. A genetically predicted increase in BMI by one standard deviation was associated with increased levels of NPNT (beta = 0.145, 95% CI: 0.084–0.206, *P* = 3.03 × 10^−6^) and HSD17B14 (beta = 0.144, 95% CI: 0.085–0.202, *P* = 1.71 × 10^−6^) using the inverse variance weighted method. MR-Egger, weighted median, and weighted mode methods yielded directionally consistent results with the inverse variant weighted method. MR: Mendelian randomization, BMI: body mass index, NPNT: nephronectin, HSD17B14: hydroxysteroid 17-beta dehydrogenase 14.

In sensitivity analyses, we tested the robustness of the MR findings with a heterogeneity test, directional pleiotropy test, and reverse causation test (see **Methods**) to only retain the proteins that were robustly influenced by BMI. We did not find significant heterogeneity for 1,304 Bonferroni-significant proteins (*I*^*2*^ < 50% for all). Of 1,304 proteins, 1,229 showed no apparent sign of directional horizontal pleiotropy with the MR Egger intercept test^30^ (*P*_Egger intercept_ > 0.05). MR analyses with the weighted median, weighted mode, and MR-Egger slope methods also showed directionally consistent results with the inverse variance weighted method for NPNT and HSD17B14 **(Figure 2** and **Supplementary Table 4)**. To assess potential reverse causation, whereby the proteins influenced BMI, we performed bidirectional MR that used protein levels as the exposures and BMI as the outcome (see **Methods**). Thirteen proteins exhibited bidirectional effects (**Supplementary Table 5**) and were excluded from further analyses.

Hence, a total of 1,216 protein levels were identified as BMI-driven proteins, which were estimated to be influenced by BMI with no apparent heterogeneity, directional pleiotropy, or reverse causation. We proceeded to the second step of our study (step 2 MR) with these 1,216 proteins, including NPNT and HSD17B14.

#### 2. BMI-driven proteins to COVID-19 severity (Step 2 MR)

Next, we evaluated the causal effects of the above-obtained BMI-driven proteins on COVID-19 outcomes, again using two-sample MR. We used cis-pQTLs for these proteins as instrumental variables and GWASs from the COVID-19 Host Genetics Initiative (release 7)^2^ as outcomes (**Supplementary Table 1)**. We used cis-pQTLs (pQTLs that reside within ± 1 Mb region around a transcription start site of a protein-coding gene) as the exposures to protect against bias from directional horizontal pleiotropy. This is because cis-pQTLs reside near the transcription start site of the protein-coding gene and are more likely to directly influence the protein levels than the trans-pQTLs^31,32^. Since cis-pQTLs are likely to directly influence the transcription or translation of their associated gene, the risk of directional horizontal pleiotropy would be greatly reduced.

We searched for the cis-pQTLs for 1,216 BMI-driven proteins using the deCODE study^26^. Following the cis-pQTL search and data harmonization, 358 and 352 proteins were tested in MR for their estimated causal effects on critically ill COVID-19 and COVID-19 hospitalization, respectively. The F-statistics for the tested proteins were all greater than 10, substantially reducing the risk of weak instrument bias^29^. F-statistics for NPNT and HSD17B14 were 252.5 and 66.3, respectively (**Supplementary Table 2**).

COVID-19 and COVID-19 hospitalization outcomes are collectively referred to as COVID-19 severity outcomes. Throughout the study, we focused on these two outcomes from the COVID-19 Host Genetics Initiative and did not include COVID-19 susceptibility outcome (reported COVID-19 infection). We did so because the determinants of COVID-19 susceptibility may reflect local testing strategy and resource allocation, which pose difficulties in the interpretation of genetic findings.

Based on a Bonferroni-adjusted threshold of *P* < 1.40 × 10^−4^, MR revealed that a one standard deviation (SD) increase in genetically predicted NPNT levels was associated with increased odds of critically ill COVID-19 (OR = 1.71, 95% CI: 1.45–2.02, *P* = 1.63 × 10^−10^) and COVID-19 hospitalization (OR = 1.36, 95% CI: 1.22–1.53, *P* = 4.52 × 10^−8^) (**Figure 3** and **Supplementary Table 6 and 7**). Similarly, a one SD increase in genetically predicted HSD17B14 levels was associated with increased odds of critically ill COVID-19 (OR = 1.92, 95% CI: 1.39–2.65, *P* = 6.85 × 10^−5^).

**Figure 3.**
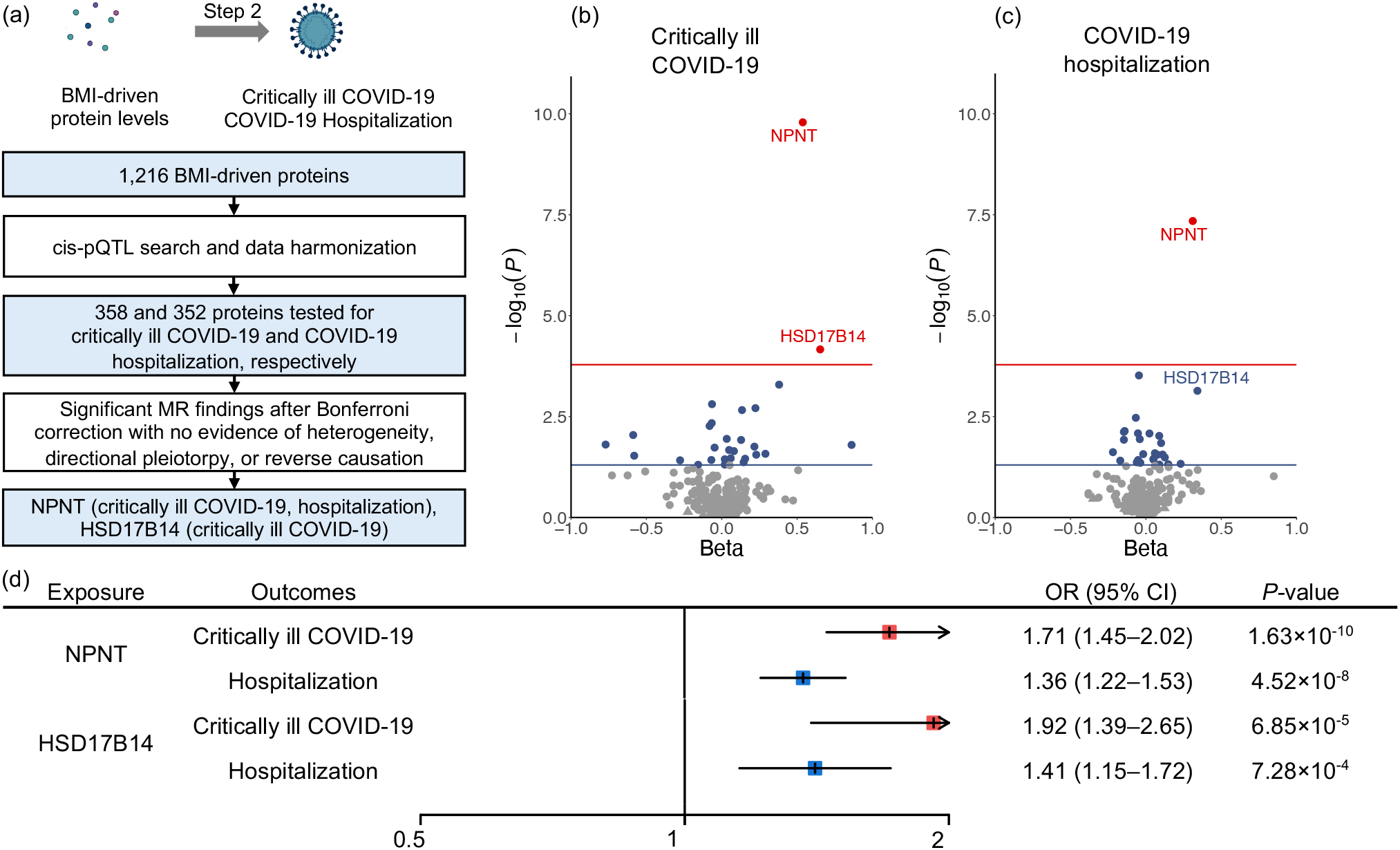
MR analyses of BMI-driven proteins on COVID-19 outcomes. (a) Flow diagram of the Step 2 MR analyses. (b, c) Volcano plot illustrating the effect of BMI on critically ill COVID-19 and (b) COVID-19 hospitalization (c) from the MR analyses using the inverse variance weighted method or Wald ratio when only one SNP was available as an instrumental variable. Red and blue horizontal lines represent *P* = 1.4 × 10^−4^ (Bonferroni correction for 358 proteins: 0.05/358) and 0.05, respectively. A proteins’ shape denotes whether the protein passed (circle) all sensitivity tests (i.e., heterogeneity, directional pleiotropy, and reverse causation assessment) or failed any of them (triangle). (d) Forest plot of the MR results for NPNT and HSD17B14, showing the odds ratio per one standard deviation increased in plasma levels of NPNT and HSD17B14 for critically ill COVID-19 and hospitalization outcomes. MR: Mendelian randomization, BMI: body mass index, NPNT: nephronectin, HSD17B14: hydroxysteroid 17-beta dehydrogenase 14, OR: odds ratio, 95% CI: 95% confidence intervals.

To verify the assumption of a lack of directional pleiotropy which can reintroduce confounding, we checked whether the cis-pQTLs for NPNT and HSD17B14 were associated with any traits or diseases using the PhenoScanner (http://www.phenoscanner.medschl.cam.ac.uk/)^33^ and Open Target Genetics (https://genetics.opentargets.org/) databases at the genome-wide significant threshold of *P* = 5 × 10^−8^. The lead cis-pQTL for NPNT (rs34712979) from the deCODE study was associated with lung-related traits (**Supplementary Table 8**). However, since NPNT has an established role as an extracellular matrix protein and fibrosis in the lungs^34-36^, it is possible that the NPNT cis-pQTL affects such traits by altering NPNT levels, and thus should not violate the assumption of no directional pleiotropy. Indeed, MR showed that NPNT levels were estimated to influence the FEV1/FVC ratio (a key lung function index used for the definition of chronic obstructive lung disease (COPD)^37^) and asthma (**Supplementary Table 9**). This suggests that these findings may be a case of vertical pleiotropy, which does not bias MR interpretation^38,39,40^. Intriguingly, the NPNT-increasing rs1662979-G allele, which increases the risk of COVID-19 severity, was found to improve lung function (i.e., higher FEV1/FVC ratio) and decrease the risk of COPD (**Supplementary Table 8 and 9**). A similar phenomenon has been reported for ELF5 and MUCB5: the COVID-19 severity risk-increasing alleles of ELF5 and MUCB5 were observed to improve lung function and decrease the risk of idiopathic pulmonary fibrosis^18^. This suggests that COVID-19 has distinct underlying mechanisms that influence the severity risk. No other cis-pQTL for NPNT or HSD17B14 was associated with any trait or disease, thereby reducing the possibility of directional pleiotropy. Next, to assess potential bias from reverse causation, we performed the MR-Steiger test, which supported a causal direction of plasma NPNT and HSD17B14 levels influencing COVID-19 severity outcomes (**Supplementary Table 10**).

#### 3. Validation analyses for NPNT and HSD17B14

##### 3.1. Step 1 MR validation

###### 3.1.1. MR analyses using body fat percentage

BMI is an easy-to-measure, widely used proxy for obesity, which can offer clinically relevant information. However, another proxy for obesity, body fat percentage, can more directly measures body fat accumulation. Thus, to evaluate whether circulating NPNT and HSD17B14 levels were influenced by body fat accumulation, we repeated step 1 MR using body fat percentage as the exposure instead of BMI. We used the same 4,907 plasma protein levels GWASs from the deCODE study^26^ as the outcomes. The F-statistic for body fat percentage for these analyses was 61.1, which indicated no evidence of weak instrument bias.

We found that one SD increase in body fat percentage was associated with increased levels of NPNT (beta = 0.14, 95% CI: 0.07–0.22, *P* = 1.23 × 10^−4,^ and HSD17B14 (beta = 0.17, 95% CI: 0.10–0.24, *P* = 3.61 × 10^−6^), (see **Methods** and **Supplementary Table 11**), consistent with the step 1 MR findings with BMI.

###### 3.1.2. Comparison with observational studies from INTERVAL

One way to assess potential biases is to test the same hypothesis using a different study design. Since each study design has its own inherent potential biases, similar results across designs can serve to strengthen causal inference through a triangulation of results^41^. Therefore, in supplementary analyses, we compared our MR findings for the effect of BMI on NPNT and HSD17B14 with published results from the INTERVAL study, which evaluated the associations between BMI and 3,622 plasma protein levels among 2,729 individuals^6^.

In the INTERVAL study, a one SD increase in BMI was cross-sectionally associated with increased NPNT (beta = 0.13, 95% CI: 0.09–0.17, *P* = 4.52 × 10^−10^), which was directionally consistent with the MR findings. On the contrary, HSD17B14 showed inconsistent results, wherein a one SD increase in BMI was not associated with HSD17B14 (beta = -0.02, 95% CI: - 0.05–0.02, *P* = 0.415).

These validation analyses collectively supported the causal effect of obesity on NPNT, but not on HSD17B14.

##### 3.2. Step 2 MR validation

###### 3.2.1. Colocalization of cis-pQTLs with COVID-19 severity outcomes

Considering that the MR analyses from Step 1 and Step 2 implicated NPNT and HSD17B14 as candidate proteins mediating the effect of BMI on COVID-19 severity, we performed colocalization to assess whether the cis-pQTLs for NPNT and HSD17B14 shared the same single causal variant with critically ill COVID-19 and hospitalization. This can test whether MR analyses were biased by LD.

The colocalization analyses revealed that NPNT had a high posterior probability of colocalization with critically ill COVID-19 and hospitalization (posterior probability (PP_shared_) > 99.9% for both) (**Figure 4**). We confirmed the robustness of the colocalization results using different priors (**Methods and Supplementary Table 12**), which consistently showed that the cis-pQTL for NPNT colocalized with COVID-19 severity outcomes. Moreover, using Combined Annotation Dependent Depletion (CADD)-scores^42^ as priors for fine-mapping, we confirmed that rs34712979, the lead cis-pQTL for NPNT, was a causal variant for critically ill COVID-19 and hospitalization with a posterior inclusion probability of > 0.99 (**Supplementary Table 13 and 14**). On the contrary, cis-pQTLs for HSD17B14 did not colocalize with either of the COVID-19 severity outcomes (**Figure 4**), indicating that the MR findings for HSD17B14 were likely to be biased by LD.

**Figure 4.**
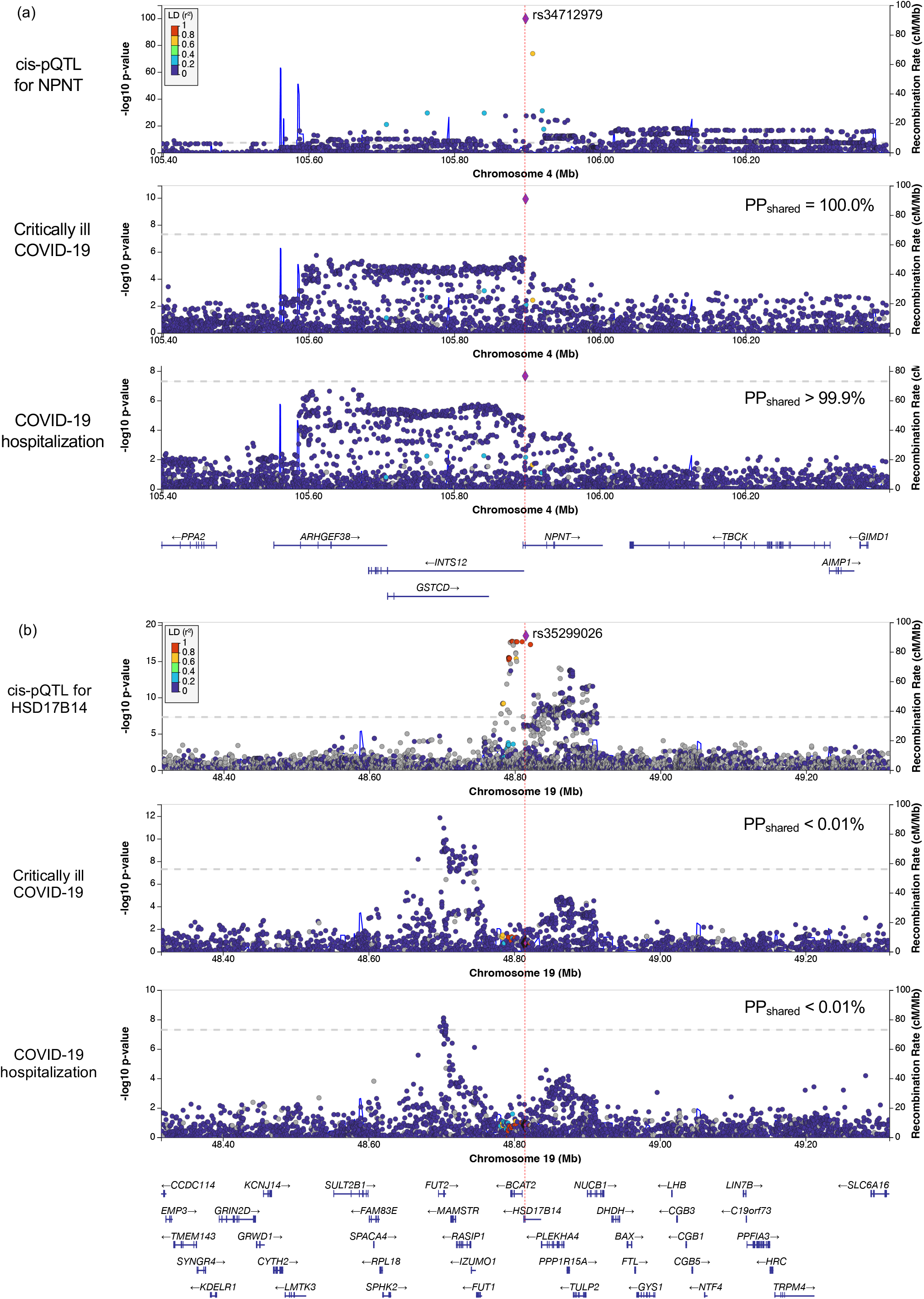
Colocalization analyses of cis-pQTL for NPNT or HSD17B14 with COVID outcomes in the 1-Mb region around rs34712979. We evaluated whether the cis-pQTL for NPNT (a) and HSD17B14 (b) shared the same causal variant with critically ill COVID-19 or COVID-19 hospitalization outcomes using colocalization. PP_shared_: Posterior probability that cis-pQTL for NPNT shares a single causal signal with the COVID-19 outcome.

###### 3.2.2. MR analyses using cis-pQTLs from different studies

To further test whether plasma levels of NPNT or HSD17B14 were causal for critically ill COVID-19, we performed additional sets of MR analyses using cis-pQTLs from different cohorts. The cis-pQTLs for NPNT and HSD17B14 were identified using data from the FENLAND study by Pietzner *et al*. (*n* = 10,708)^27^ and the AGES Reykjavik study by Emilsson *et al*. (*n* = 3,200)^28^. We assessed the PhenoScanner and Open Target Genetics databases for additional associations for these cis-pQTLs with other phenotypes and found none.

MR analyses using these cis-pQTLs estimated a consistent causal effect of NPNT on critically ill COVID-19 (the FENLAND study: OR = 1.89, 95% CI: 1.56–2.29, *P* = 1.21 × 10^−10^; the AGES Reykjavik study: OR = 1.25, 95% CI: 1.06–1.48, *P* = 8.26 × 10^−3^) and COVID-19 hospitalization (the FENLAND study, OR =1.45, 95% CI: 1.28–1.66, *P* = 2.17× 10^−8^; the AGES Reykjavik study, OR = 1.17, 95% CI: 1.04–1.31, *P* = 7.30 × 10^−3^).

In contrast, the estimated causal effects of HSD17B14 on these COVID-19 outcomes were not supported (**Supplementary Table 15**). Given the lack of colocalization and replication, we concluded that the initial finding indicating a causal effect of HSD17B14 on COVID-19 severity outcomes using a cis-pQTL from the deCODE study was likely biased by a difference in LD structure. Therefore, we excluded HSD17B14 from further evaluation.

###### 3.2.3. Comparing with observational associations using BQC19

Considering that the MR analyses (Step 2 MR) used plasma protein levels in a non-infectious state from the deCODE study as the exposures, in supplementary analyses, we observationally assessed whether the plasma levels of NPNT in a non-infectious state were associated with the risk of COVID-19 using logistic regression in 293 individuals from the BQC19 cohort. We restricted the analyses to individuals who met our criteria of critically ill COVID-19, COVID-19 hospitalization, or COVID-19 negative controls (see **Methods)**.

Logistic regression analysis adjusting for age, sex, and sample collection batch showed that increased non-infectious plasma levels of NPNT were associated with the increased risk of critically ill COVID-19 (OR = 1.47, 95% CI: 1.05–2.06, *P* = 2.60 × 10^−2^) and COVID-19 hospitalization (OR = 1.49, 95% CI: 1.11–2.02, *P* = 9.87 × 10^−3^), which were directionally consistent with the step 2 MR findings (**Supplementary Table 16**).

#### 4. Follow-up analyses for the putatively causal protein (NPNT)

##### 4.1. Colocalization of NPNT’s cis-pQTL with eQTL and sQTL

The observed differences in NPNT levels measured by the SomaScan assay may be explained by levels of a particular isoform, rather than total NPNT levels. Thus, we performed colocalization analyses to evaluate whether the cis-pQTL share the same causal variant with either the total RNA expression level or a specific isoform level of NPNT. Given that the lung is a primary target organ in the context of COVID-19 severity, NPNT is highly expressed in the lung, NPNT splice isoforms have previously been implicated as a risk factor for COPD and other lung outcomes^43,44,45^, and SNPs influencing a specific isoform level would be sQTLs, we performed colocalization analysis of the cis-pQTL with eQTLs and sQTLs for NPNT in lung tissue from GTEx^46^.

Colocalization analysis of NPNT pQTL and sQTL within a one-megabase (1-Mb) region (±500 kb) surrounding the lead cis-pQTL (rs34712979) revealed that there was a high probability of pQTL and sQTL sharing a single causal variant (posterior probability for a shared causal signal (PP_shared_) = 100.0%). However, this was not true for the eQTL (PP_shared_ < 0.01%; **Figure 5**). We confirmed the robustness of the colocalization results using different priors (**Methods and Supplementary Table 17**).

**Figure 5.**
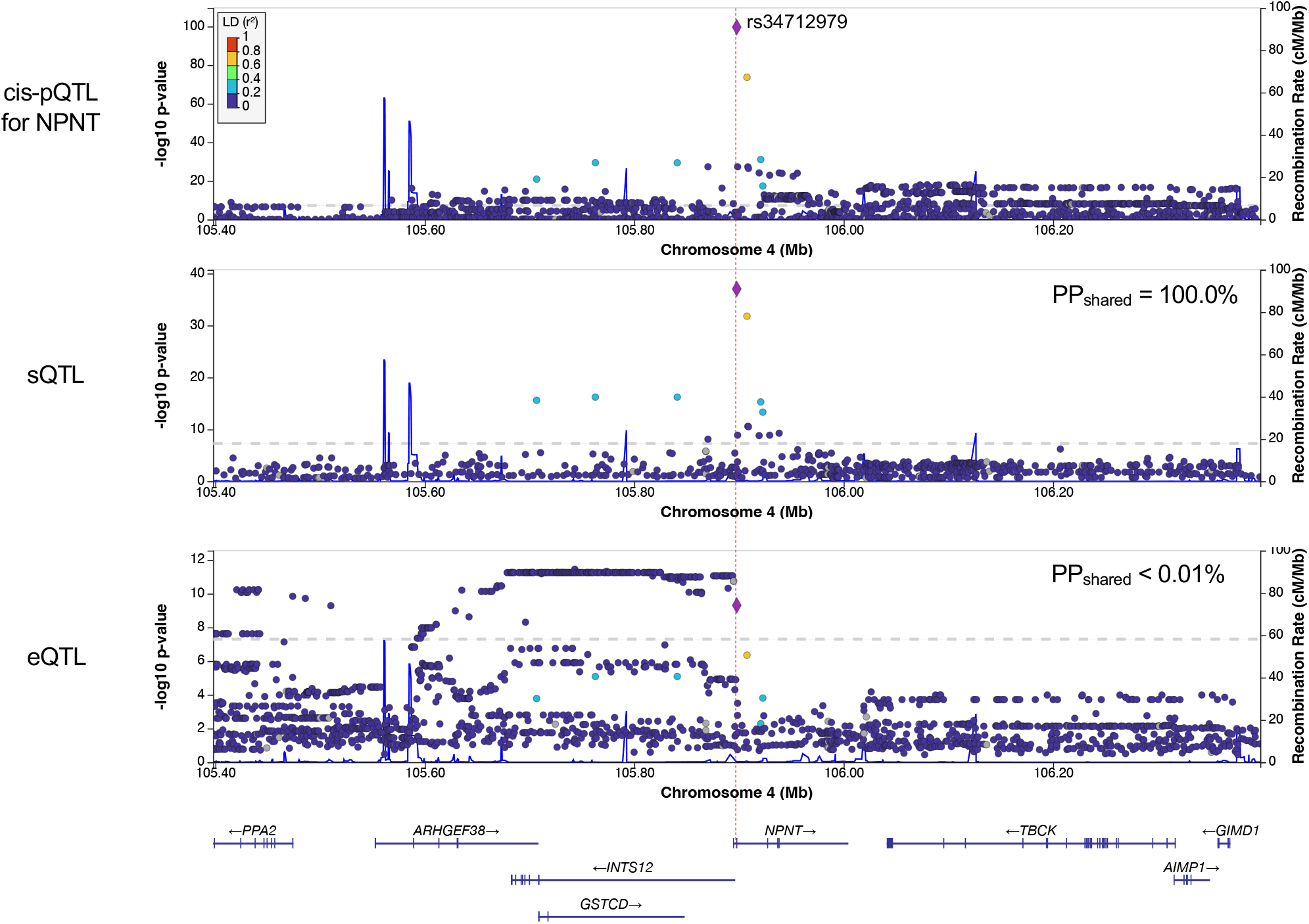
Colocalization analyses of cis-pQTL with sQTL and eQTL for NPNT. PP_shared_: Posterior probability that the cis-pQTL for NPNT shared a single causal signal with its sQTLs or eQTLs in the lung.

Given that the thyroid, lung, and arteries are the top three *NPNT*-expressing tissues according to the GTEx^46^, we performed the same colocalization analyses in the thyroid and arteries (aorta, tibial, and coronary) and found that the pQTL of NPNT also colocalized with sQTL in these tissues (PP_shared_ = 100.0% for all). Furthermore, it colocalized with the eQTL in the aorta and tibial artery (PP_shared_ = 100.0% for both), but not in the thyroid or coronary artery (PP_shared_ = 11.9% and 5.8%, respectively).

Notably, the lead cis-pQTL from the deCODE study, i.e., rs34712979, was also identified by the FENLAND study^27^ and has been reported to create a cryptic splice acceptor site, which inserts a three-nucleotide sequence coding a serine residue at the 5’-splice site of exon 2, resulting in perturbations of the alpha-helix motif^43^. Further, this lead cis-pQTL (rs34712979) and another cis-pQTL (rs78213340) from the AGES Reykjavik study—that was not in high LD with rs34712979 (*r*^*2*^ = 0.234)—were both associated with the same exon-skipping splicing in the lung in GTEx (**Supplementary Table 18**). Hence, two different cis-pQTLs of NPNT from three different studies—all using SomaScan—were associated with the same splicing pattern, suggesting that the SomaScan assay measures a specific isoform of the NPNT protein.

Collectively, these findings indicate that the SomaScan NPNT-targeting aptamer measures specific isoform levels and the specific isoform of NPNT with a serine insertion at the N-terminus, influences the effect of NPNT on COVID-19 severity.

##### 4.2. Single-cell RNA-sequencing data of SARS-CoV-2-infcted lungs

To gain insights into the biological role of NPNT in SARS-CoV-2-infected lungs, we explored the lung cell types that significantly expressed the *NPNT* gene by analyzing single-cell RNA-sequencing data from lung autopsy samples (106,792 cells) of 16 patients who died of COVID-19^47^ (Single Cell Portal of the Broad Institute (Accession ID: SCP1052)).

We found that *NPNT* is widely expressed in lung cell types, including epithelial cells (type 1 and type 2 alveolar cells) and fibroblasts, highlighting its role in air exchange and fibrosis. Furthermore, we conducted a subgroup analysis of SARS-CoV-2-positive and -negative cells and found that *NPNT* was significantly expressed in SARS-CoV-2-positive alveolar cells and fibroblasts (permutation test *P* < 0.001, see **Methods** for details) (**Figure 6**).

**Figure 6.**
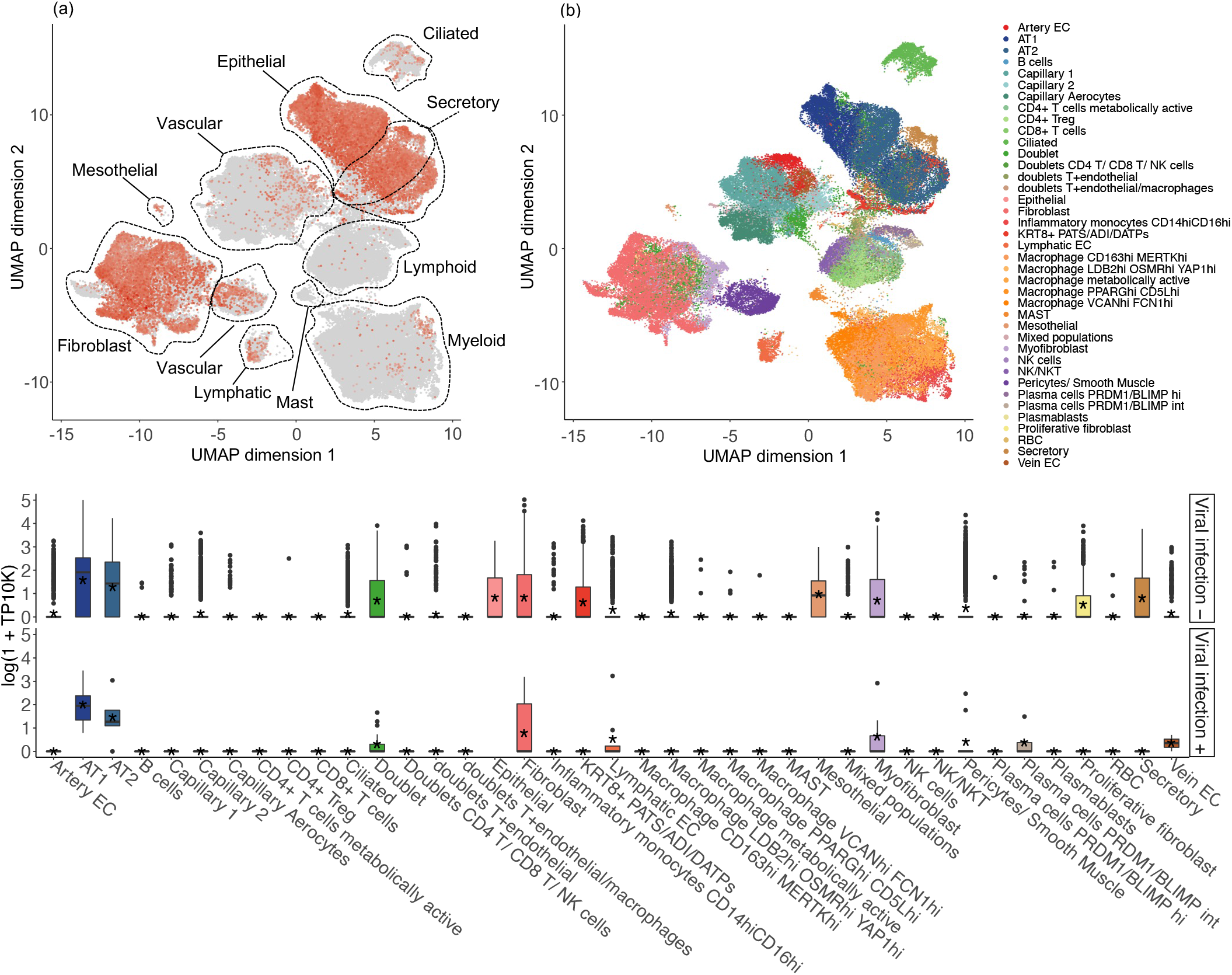
NPNT expression levels in lung cell types from COVID-19 lung autopsy samples at single-cell resolution. (a) NPNT expression levels of each cell type at single-cell resolution in the 16 lung donors with COVID-19. (b) Twenty-eight annotated cell types of the lung. (c) NPNT expression status in 106,449 SARS-CoV-2 non-infected cells (viral infection −, top panel) or 343 SARS-CoV-2 infected cells (viral infection +, bottom panel) in 16 lung donors. NPNT expression levels of 28 cell types in the two groups are shown in a box plot. In each box, the horizontal line denotes a median value of the expression levels, and the asterisk inside each box denotes the mean value. Each box extends from the 25th to the 75th percentile of each group. Whiskers extend 1.5 times the interquartile range from the top and bottom of the box. Log (TP10K+1) was calculated by normalizing original gene counts by total unique molecular identifiers (UMI) counts, multiplying by 10,000 (TP10K), and then taking the natural logarithm.

##### 4.3. Mediation analysis

Since BMI has likely thousands of effects upon human physiology and creates an important perturbation of the proteome, we wanted to estimate the proportion of the effect of BMI that was mediated only through plasma NPNT levels. To do so, we performed a mediation analysis using network MR with the product of coefficients method to understand the extent to which plasma NPNT levels mediate the association between BMI and critically ill COVID-19 and COVID-19 hospitalization.

For critically ill COVID-19, we estimated the effect of BMI on critically ill COVID-19 mediated by plasma NPNT levels (**Figure 7**). We first estimated the effect of BMI on plasma NPNT levels and then multiplied this estimate by the effect of plasma NPNT levels on critically ill COVID-19 (see **Methods** for further details). The ratio of the effect mediated by NPNT was calculated by dividing the NPNT-mediated effect estimate by the total effect estimate of BMI on critically ill COVID-19. We repeated the same process for COVID-19 hospitalization.

**Figure 7.**
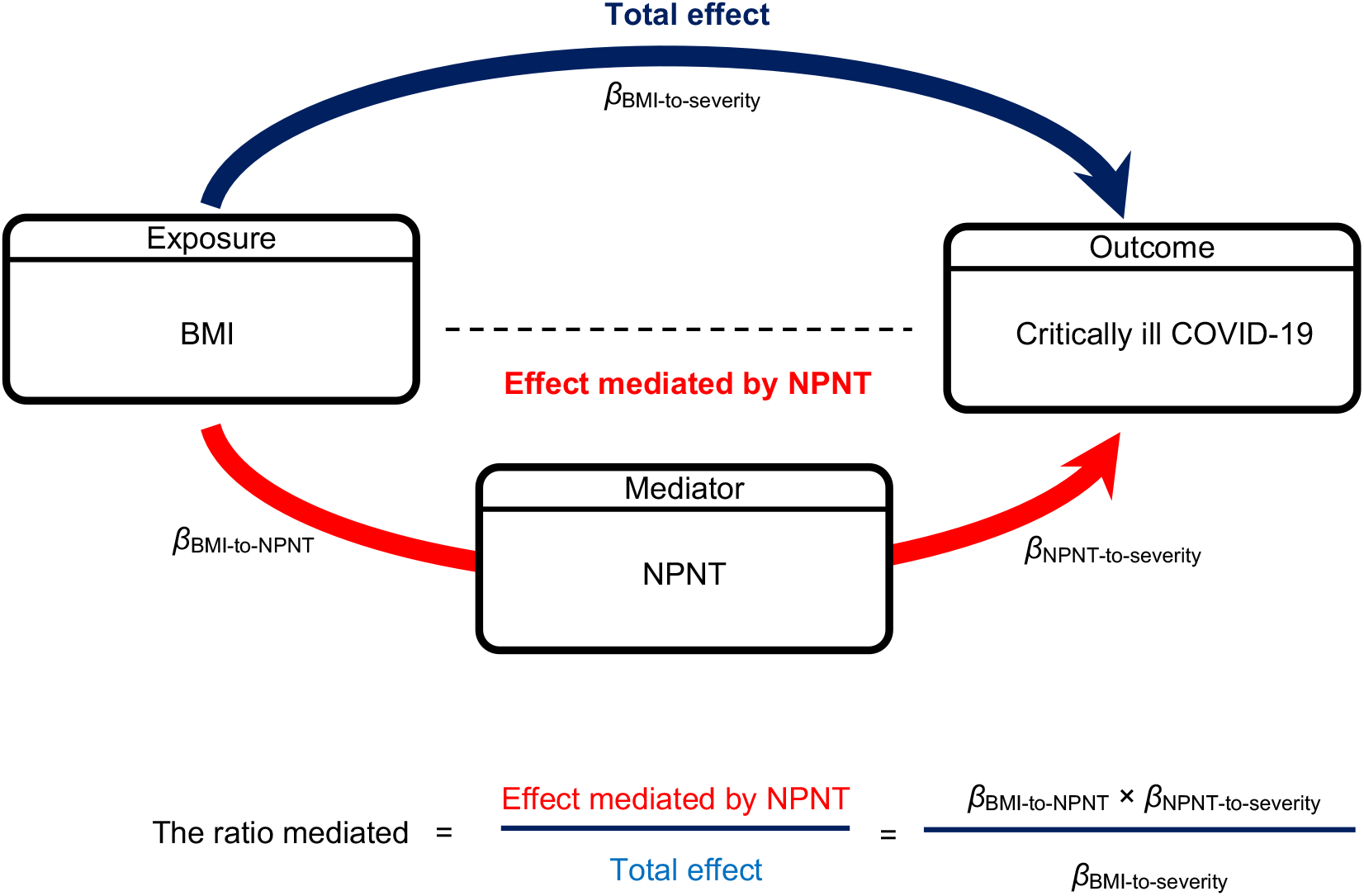
MR mediation analysis illustrated by the directed acyclic graph. The dark blue arrow represents the total effect of BMI on critically ill COVID-19. The red arrow represents the effect of BMI on critically ill COVID-19 mediated by NPNT. For the effect of BMI on critically ill COVID-19 mediated by NPNT, the product of coefficients method calculates the proportion mediated by multiplying β_BMI-to-NPNT_ and β_NPNT-to-severity_, where β_BMI-to-NPNT_ is the effect of BMI on NPNT and β_NPNT-to-severity_ is the effect of NPNT on critically ill COVID-19. We evaluated the proportion mediated for the effect of obesity-related exposures (i.e., BMI, body fat percentage, and body fat mass) on COVID-19 severity outcomes (i.e., critically ill COVID-19 and COVID-19 hospitalization). BMI: Body mass index; MR: Mendelian randomization; NPNT: nephronectin.

We found that plasma NPNT levels partially mediated the total effect of BMI on critically ill COVID-19 (proportion mediated = 13.9%, 95% CI: 6.1–21.6%, *P* = 4.52 × 10^−4^) and COVID-19 hospitalization (proportion mediated = 10.6%, 95% CI: 4.4–16.7%, *P* = 8.02 × 10^−4^) (**Supplementary Table 19**).

In supplementary analyses, we evaluated whether NPNT mediates the total effect of body fat percentage on the COVID-19 severity outcomes. We found that plasma NPNT levels mediated the total effect of body fat percentage on critically ill COVID-19 (proportion mediated = 9.5%, 95% CI: 3.6–15.4%, *P* = 1.59 × 10^−3^) and COVID-19 hospitalization (proportion mediated = 7.7%, 95% CI: 2.7–12.7%, *P* = 2.57 × 10^−3^). We also found consistent results for body fat mass; plasma NPNT levels mediated the total effect of body fat mass on critically ill COVID-19 (proportion mediated = 13.4%, 95% CI: 6.1–20.6%, *P* = 2.85 × 10^−4^) and COVID-19 hospitalization (proportion mediated = 9.9%, 95% CI: 4.3–15.6%, *P* = 5.61 × 10^−4^) (**Supplementary Table 19**). All of these results consistently suggested that plasma NPNT levels partially mediated the effect of obesity, measured by BMI, body fat percentage, or body fat mass, on COVID-19 severity outcomes.

Throughout the above analyses, we did not adjust the exposure while estimating the effect of the mediator on the outcome (β_NPNT-to-severity_ in **Figure 7**) to avoid weak instrument bias (see **Methods**). This approach was also used in previous studies^48,49,50^. In supplementary mediation analyses, we found that adjusting for the exposure when estimating β_NPNT-to-severity_ (i.e., Sobel test) also support the role of NPNT as a mediator for the effect of obesity-related exposures (BMI, body fat percentage, and body fat mass) on COVID-19 severity outcomes; however, the estimated proportion mediated was modest, likely due to weak instrument bias (**Supplementary Table 20**).

##### 4.4. Multivariable MR analyses of body fat and fat-free mass

Given the consistent evidence that BMI influenced NPNT levels, which in turn influence COVID-19 severity, we aimed to identify a way of modulating plasma NPNT levels by gaining a better understanding of how the estimated causal effect of BMI on plasma NPNT levels was influenced by fat or fat-free mass in humans. In the Step 1 MR, we showed that increased BMI is estimated to increase plasma levels of NPNT. However, BMI is a function of height and weight and does not take into account body compositions, such as body fat and fat-free mass. Thus, we specifically assessed their independent effects on plasma NPNT levels. For this, we performed multivariable MR using body fat and fat-free mass as the exposures and plasma NPNT levels as the outcomes (see **Methods**). Conditional F-statistics for body fat mass and fat-free mass were 38.8 and 59.3, respectively, which did not indicate weak instrument bias (which is suspected when F-statistics is less than 10) (**Supplementary Table 21**)^51^.

Multivariable MR using inverse variance weighted method found that a one SD increase in body fat mass was associated with increased plasma levels of NPNT (beta = 0.21, 95% CI: 0.14–0.28, *P* = 2.74 × 10^−9^), whereas a one SD increase in body fat-free mass was associated with decreased plasma levels of NPNT (beta = -0.13, 95% CI: -0.22, -0.05, *P* = 2.98 × 10^−3^) (**Figure 8**). In sensitivity analyses, Q-statistics for instrumental validity did not suggest evidence of pleiotropy (Q-statistics = 940.8, *P*_Q-statistics_ = 0.09). Multivariable MR-Egger also showed directionally consistent results, and no evidence of directional pleiotropy was observed with the MR-Egger intercept test (**Supplementary Table 21**).

**Figure 8.**
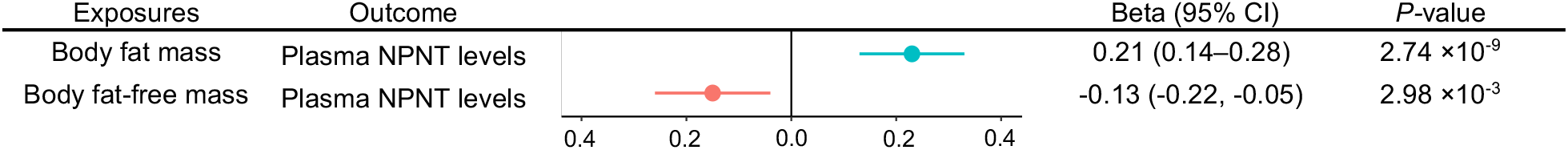
Multivariable MR analysis for evaluating independent effects of body fat and fat-free mass on plasma NPNT levels. We performed multivariable MR with the inverse variance weighted method using body fat and fat-free mass as the exposures and plasma NPNT levels as the outcome. MR: Mendelian randomization; NPNT: nephronectin.

Further, to test the influence of body fat and fat-free mass on COVID-19 severity outcomes, we conducted multivariable MR analyses using body fat mass and fat-free mass as the exposures and either critically ill COVID-19 or COVID-19 hospitalization as the outcome. We found that a one SD increase in body fat mass was associated with increased odds of critically ill COVID-19 (OR = 1.89, 95% CI: 1.65–2.16, *P* = 4.83 × 10^−20^) and COVID-19 hospitalization (OR = 1.59, 95% CI: 1.45–1.75, *P* = 4.28 × 10^−22^), whereas a one SD increase in body fat-free mass was associated with a decreased risk of critically ill COVID-19 (OR = 0.77, 95% CI: 0.65– 0.91, *P* = 1.94 × 10^−3^) and COVID-19 hospitalization (OR = 0.87, 95% CI: 0.77–0.97, *P* = 1.57 × 10^−2^). Q-statistics for instrumental validity and the MR-Egger intercept test did not show evidence of pleiotropy (**Supplementary Table 21**).

These findings suggest that decreasing body fat mass and increasing fat-free mass (e.g., through actions such as appropriate diet and exercise) can reduce plasma NPNT levels, and thus reduce the risk of critically ill COVID-19, thereby indicating NPNT as an actionable target.

## Discussion

In the present study, we conducted MR analyses and found that NPNT likely mediates an important proportion of the effect of obesity on COVID-19 severity. Considering that BMI is a highly polygenic trait with more than 530 associated loci^25^ and influences more than 1,200 circulating proteins, as shown in the present and previous studies^6,7^, it is remarkable that a single protein explains a reasonably large proportion of the effect of BMI and other obesity-related traits on COVID-19 severity outcomes. Additionally, colocalization analyses provided evidence that the NPNT cis-pQTL is shared with the lung sQTL for NPNT, which leads to a splice isoform with a serine insertion at the N-terminus of NPNT^43^. These results suggest that NPNT mediates a proportion of obesity’s effect on critically ill COVID-19 and that this effect may be conferred by alternative splicing of *NPNT* which introduces a serine residue at its N-terminus.

NPNT, or nephronectin, is an extracellular matrix protein that controls integrin binding activity. It is known as a functional ligand of integrin α8/β-1 in kidney development and is also associated with the development, remodeling, and survival of various tissues through the binding of integrins^52,53^. NPNT has also been implicated in inflammation and autoimmunity^54,55^, which may align with the suggested role of obesity-induced inflammation in COVID-19 severity^56^. In addition, *NPNT* is expressed in lung alveolar cells and fibroblasts^57,58^, and recent GWASs have identified the same NPNT splice variant, rs34712979, to be associated with lung function-related traits (e.g., forced expiratory volume (FEV1), forced vital capacity (FVC), and FEV1/FVC ratio) and COPD^44,45^. Our single-cell RNA-sequencing analysis on COVID-19 lung autopsy samples also found that *NPNT* was expressed in SARS-CoV-2-infected alveolar cells and fibroblasts, suggesting its role in air exchange and fibrosis in SARS-CoV-2-affected lungs. These findings collectively suggest that NPNT alternative splicing, which results in an isoform with a serine insertion in the N-terminus of the protein, increases the risk of deterioration of lung function and lung diseases such as COPD and COVID-19. Future studies are required to investigate the functional properties of this alternative splice isoform.

Our findings may have important clinical implications. The multivariable MR approach showed that decreasing body fat mass and increasing body fat-free mass, which can be achieved through non-pharmacological interventions such as exercise and appropriate diet^59^, can reduce plasma NPNT levels and the risk of COVID-19 severity outcomes. Moreover, recent trials have shown GLP-1/GIP co-agonist tirzepatide^60,61^ and GLP-1 receptor agonists including semaglutide^62,63^ and liraglutide^64,65^ can reduce body fat mass, while preserving body fat-free mass. We believe that these findings are important because they offer the possibility to potentially modulate plasma NPNT levels, using available therapies. Such hypotheses require further investigations in clinical trials.

This study has both strengths and weaknesses. MR and colocalization analyses robustly implicated NPNT as a causal mediator of the relationship between BMI and COVID-19 severity. The robustness of the MR findings was enhanced by the large sample sizes used to derive the findings. To the best of our knowledge, this is the first study to identify a mediator of obesity on COVID-19 employing a two-step MR approach, although a previous study using a similar framework with limited statistical power earlier in the pandemic could not identify a strong protein signal^66^. Furthermore, our MR findings withstood multiple sensitivity analyses and were supported by colocalization, fine-mapping, replication MR, observational evaluation, and RNA-sequencing studies in COVID-19 lung samples. Intriguingly, multivariable MR showed that plasma NPNT levels were increased by body fat mass but decreased by fat-free mass, indicating that the effect of BMI on plasma NPNT levels was driven by body fat mass and partially counteracted by fat-free mass. These findings suggest that NPNT could be a potential intervention target in individuals with obesity to prevent critically ill COVID-19 and highlight one of the possible mechanisms by which appropriate diet and exercise can confer risk reduction of COVID-19 severity through the reduction in plasma NPNT levels.

This study also has important limitations. First, MR and colocalization analyses were restricted to individuals of European ancestry to avoid confounding by population stratification and heterogeneity of genetic associations across different ancestries. Whether NPNT mediates the effect of BMI on COVID-19 severity in populations of non-European ancestries requires further investigation. Second, we did not perform sex-stratified analysis due to the unavailability of sex-specific datasets. Third, there were no genome-wide significant genetic instrumental variables for BMI in the 1-Mb region around the cis-region of NPNT (1-Mb around the transcription start site), we could not pinpoint a single genetic variant that directly links BMI to NPNT. Hence, the effect of BMI on NPNT is likely mediated by a combination of multiple trans-effects, which was also indicated by the scatter plot in the MR analysis for NPNT. However, this is not surprising considering that BMI is a highly polygenic, complex trait, and multiple pathways could confer the effect of BMI on other diseases^67^. Fourth, we do not know the molecular mechanism by which BMI influences the splice isoform. Previous studies have suggested that obesity is associated with alternative splicing^68-70^ and extracellular matrix protein remodeling^71^, which may align with our findings. However, further investigation is required to clarify the specific molecular mechanisms involved. Lastly, we do not rule out the possibility that total levels of NPNT (all isoforms) measured by the NPNT-targeting aptamer mediate the effect of obesity on COVID-19 severity. However, given the evidence provided by the MR, colocalization, fine-mapping, and a biological understanding of lead cis-pQTL (i.e., the variant causes alternative splicing resulting in perturbations of the alpha-helix motif in the lung), it is likely that the specific isoform measured by the aptamer is driving the effect. Nevertheless, isoform-specific measurements (e.g., mass spectrometry) will be required to confirm these findings.

In conclusion, we integrated a two-step MR approach, sensitivity analyses, colocalization, fine-mapping, single-cell RNA-sequencing, and mediation analyses to identify NPNT as an important mediator of the effect of obesity on COVID-19 severity outcomes. We also showed that decreasing body fat mass and increasing fat-free mass (e.g., by actions such as exercise and appropriate diet) can lower NPNT levels, and thus may improve COVID-19 severity outcomes. These findings provide actionable insights into how obesity influences COVID-19 severity.

## Methods

### Step 1: BMI to plasma proteins

#### Two-sample MR

##### BMI GWAS

We used the BMI GWAS meta-analysis with the largest sample size, comprising 693,529 European ancestry individuals from the GIANT consortium and UK Biobank^25^. The consortium details are provided in **Supplementary Table 1**, and post-hoc power calculation is provided in **Supplementary Table 22**.

##### Proteomic GWAS

For GWAS of plasma protein levels, we used the largest proteomic GWAS available^26^, which measured 4,907 proteins in 35,559 individuals of European ancestry using the SomaLogic SomaScan assay v4 (SOMAscan, SomaLogic, Boulder, Colorado, USA).

##### Two-sample MR

The effect of BMI on plasma protein levels was assessed using the inverse variance weighted method with a random-effects model in TwoSampleMR v.0.5.6^72^ (https://mrcieu.github.io/TwoSampleMR/). The instrumental variables for the exposure were defined as genome-wide significant and independent single nucleotide polymorphisms (SNPs) (*P* < 5 × 10^−8^; *r*^*2*^ < 0.001, with a clumping window of 10 Mb). SNPs in the human major histocompatibility complex (MHC) region at chromosome 6: 28,477,797–33,448,354 (GRCh37) were excluded considering its complex LD structure. We used PLINK v1.9^73^ (http://pngu.mgh.harvard.edu/purcell/plink/) to obtain instrumental variables by clumping SNPs using the 1000 Genomes Project European reference panel^74^ and applying an LD threshold of *r*^*2*^ < 0.001. The genome-wide significant independent SNPs with the lowest *P*-value were selected from each LD block. If instrumental variable SNPs were not present in an outcome GWAS, we used proxy SNPs (*r*^*2*^ > 0.8 with the original SNP). Proxy SNPs were identified using snappy v1.0 (https://gitlab.com/richards-lab/vince.forgetta/snappy) with 1000 Genomes Project’s European reference panel^74^. Data harmonization and MR analyses were conducted using TwoSampleMR v0.5.6. Specifically, data harmonization was performed with the “harmonise_data()” function using default settings, including the removal of palindromic SNPs that have minor allele frequency above 0.42. MR was performed using the “mr()” function. A Bonferroni correction was used to set a statistical significance threshold by dividing 0.05 by the number of proteins (*P* = 1.0 × 10^−5^ (0.05/4907)). We note that this correction is overly-conservative since many proteins are non-independent. We did so to safeguard against false positive findings. In a heterogeneity test, we calculated *I*^*2*^ statistics using “Isq()” function and heterogeneity *P*-value with “mr_heterogeneity()” function; results with an *I*^*2*^ > 50% and heterogeneity *P*-value (Q_pval) < 0.05 were considered to be heterogeneous (substantial heterogeneity)^75^. For evaluating directional pleiotropy, we used the MR-Egger intercept test, which was performed using the “mr_pleiotropy_test()” function; where directional pleiotropy was considered to be present when the MR-Egger intercept differed from the null (*P* < 0.05). We note that even in the presence of moderate heterogeneity, balanced horizontal pleiotropic effects would not violate the MR assumption of a lack of directional pleiotropy^76,77^. For NPNT and HSD17B14, we used MR-Egger, weighted median, and weighted mode methods as additional sensitivity analyses to evaluate the directional consistency of beta coefficients with the inverse variance weighted method.

For reverse MR, whereby the effects of plasma protein levels on BMI were assessed, we used cis-pQTLs from the deCODE study as the exposures and BMI GWAS as the outcome, deploying the inverse variance weighted method or Wald ratio method when only one SNP was available as an instrumental variable. After the cis-pQTL search, proxy, and data harmonization, 357 proteins were tested in MR for their estimated reverse effects. Results with *P* < 1.4 × 10^−4^ (0.05/357; Bonferroni correction) were considered statistically significant.

We used BMI GWAS from the UK Biobank (not the meta-analysis GWAS) because multiple variants, including cis-pQTL for NPNT from the deCODE study (rs34712979), were dropped during the stringent quality control process of the meta-analysis (e.g., rs34712979 was not present in the GIANT GWAS and thus dropped during the meta-analysis process).

To assess statistical power, F-statistics were calculated as previously described^78^ using the following formula: 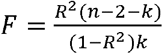 where: *R*^*2*^ = proportion of variance in the exposure trait and *k* = number of instrumental variables (**Supplementary Table 2)**. We also performed post-hoc power calculations using an online power calculator for Mendelian randomization (https://sb452.shinyapps.io/power/) (**Supplementary Table 22**).

### Step 2: BMI-driven proteins to COVID-19 severity outcomes

#### Two-sample MR

##### Cis-pQTL GWAS

We identified cis-pQTLs from the GWAS of 4,907 proteins in 35,559 individuals of European ancestry from the deCODE study^26^. Cis-pQTLs were defined as pQTLs located within 1-Mb around a transcription start site of a protein-coding gene. Further details of the dataset are provided in **Supplementary Table 1**. Genetic coordinates of transcription start sites of each gene used to define cis-pQTLs are provided in Supplementary Table 2 of the deCODE study^26^.

##### COVID-19 severity outcome GWAS

For COVID-19 outcomes, we used a GWAS meta-analysis from the COVID-19 Host Genetics Initiatives data release 7 (https://www.covid19hg.org/). The outcomes included critically ill COVID-19 (13,769 cases and 1,072,442 controls) and COVID-19 hospitalization (32,519 cases and 2,062,805 controls). These two outcomes were collectively referred to as COVID-19 severity outcomes. Critically ill COVID-19 was defined as a requirement for respiratory suport among hospitalized individuals with laboratory-confirmed SARS-CoV-2 infection or death due to COVID-19. COVID-19 hospitalization was defined as a laboratory-confirmed SARS-CoV-2 infection that required hospitalization.

##### Two-sample MR

Using the data above, we carried out two-sample MR for the effects of plasma protein levels on COVID-19 severity outcomes. We used the inverse variance weighted method for proteins with ≥2 instrumental variables and the Wald ratio method for proteins with a single instrumental variable. When an instrumental variable SNP was not found in an outcome GWAS, we searched and used a proxy SNP (*r*^*2*^ < 0.8 with the original SNP) using snappy v1.0 (https://gitlab.com/richards-lab/vince.forgetta/snappy) with 1000 Genomes Project European reference panel^74^. Results with a *P* < 1.4 × 10^−4^ (0.05/358; Bonferroni correction with the number of proteins tested in the Step 2 MR) were considered statistically significant. We used the MR-Egger intercept test to assess lack of directional pleiotropy for proteins with three or more genetic instrumental variables. However, we could not use this test for NPNT because it had fewer than three genetic instrumental variables. Therefore, we used the PhenoScanner (http://www.phenoscanner.medschl.cam.ac.uk/) and Open Targets Genetics (https://genetics.opentargets.org/) databases to test whether variants had potential pleiotropic associations with other diseases or traits. Associations with *P* < 5 × 10^−8^ were considered statistically significant. We also assessed reverse causation, wherein the effect of COVID-19 severity may influence plasma protein levels with the MR-Steiger test using “directionality_test()” function from TwoSampleMR v.0.5.6^72^.

### 3. Validation analyses for proteins prioritized by step 1 and step 2 MR (NPNT and HSD17B14)

#### 3.1. Step 1 MR validation

##### 3.1.1. MR analysis using body fat percentage

We repeated the step 1 MR (described above) for NPNT and HSD17B14 using body fat percentage as the exposure instead of BMI. For this, we used body fat percentage GWAS in 454,633 individuals of European ancestry from the UK Biobank, obtained from the IEU OpenGWAS project (https://gwas.mrcieu.ac.uk/). The accession ID was ukb-b-8909.

##### 3.1.2. Comparison of the MR findings with the published observational association study from INTERVAL

The INTERVAL study was a prospective cohort study conducted in England. The study recruited ∼50,000 participants of primarily European ancestry without a self-reported history of any major disease. The recruitment took place between 2012 and 2014, before the COVID-19 pandemic. The study measured 3,622 plasma protein levels in 2,737 individuals using the SomaScan assay. The study performed a linear regression to evaluate associations between BMI and plasma protein levels, adjusting for age and sex^6^. The derived beta estimates represented normalized SD-unit difference in each protein level per one SD (4.8 kg/m^2^) increase in BMI. Further details of the study have been described in depth previously^6^.

#### 3.2. Step 2 MR validation

##### 3.2.1. Colocalization of cis-pQTLs with COVID-19 severity outcomes

To identify potential bias caused by confounding owing to LD, we performed colocalization to evaluate whether cis-pQTLs shared a single causal variant between the three COVID-19 outcomes for the causal proteins and NPNT or HSD17B14. We used the coloc R package v5.1.0^79^ (https://chr1swallace.github.io/coloc/) to evaluate all SNPs within the 1-Mb region around the top cis-pQTL for the NPNT (defined as the cis-pQTL with the smallest *P*-value). The posterior probability of hypothesis 4 (H_4_) or PP_shared_ (two traits sharing a single causal variant) > 0.8 was considered to indicate strong evidence of colocalization. Colocalization analyses were conducted using the coloc R package v5.1.0^79^, using default priors of p1 = p2 = 10^−4^ and p12 = 10^−5^, where p1 is a prior probability that only trait 1 has a genetic association in the region, p2 is a prior probability that only trait 2 has a genetic association in the region, and p12 is a prior probability that both trait 1 and trait 2 share the same genetic association in the region. To test the robustness of the results, we evaluated different combinations of priors: p1 = c(10^−4^, 10^−5^, 10^−6^), p2 = c(10^−4^, 10^−5^, 10^−6^), p12 = c(10^−5^, 5 × 10^−6^, 10^−6^), as performed previously^18^.

###### Fine-mapping of the NPNT region in the COVID-19 severity GWAS

The COVID-19 Host Genetics Initiatives release 7 data for those of European ancestry with critically ill COVID-19 and hospitalization was inputted into FINEMAP v1.4^80^ (http://www.christianbenner.com/) using default options. We assumed a maximum of one causal SNP, given a single GWAS significant SNP in the NPNT region. Summary statistics were filtered for SNPs present in the European ancestry, as done by Huffman *et al*^81^. We set prior probabilities using scaled CADD-PHRED scores^42^ (https://cadd.gs.washington.edu/). The scaled CADD-PHRED scores were normalized so that their sum equaled one. The variance of effect size was set such that the maximum odds ratio a variant can have with 95% probability is two, then scaled using the case-control ratio as described previously^81^.

##### 3.2.2. MR analyses using cis-pQTLs for NPNT and HSD17B14 from different cohorts

We also obtained cis-pQTLs and their beta estimates for plasma protein levels of NPNT and HSD17B14 from the FENLAND study and AGES Reykjavik study (**Supplementary Table 10**). We repeated the two-sample MR using these cis-pQTLs as instrumental variables and the COVID-19 severity outcomes as described above.

##### 3.2.3. Comparing with observational associations using BQC19

###### The BQC-19 cohort

BQC19 (Biobanque Québécoise de la COVID-19) is a province-wide biobank that provides global access to important biological and clinical data from patients with COVID-19 and control subjects in Québec, Canada (https://www.bqc19.ca/). Blood samples were collected in acid citrate dextrose (ACD) tubes from 264 SARS-CoV-2 infectious and 463 non-infectious patients of European ancestry (see **Supplementary Information** for the definition of infectious or non-infectious state). Detailed description of sample processing can be found in the **Supplementary Information**. Briefly, the samples underwent proteomic profiling on the SomaScan v4 assay, and 4,907 aptamers were used for analysis, consistent with the deCODE study^26^. For quality control, protein levels were natural log-transformed and then batch-corrected using the ComBat function implemented in the sva R package v3.44.0^82^.

###### Logistic regression analysis in BQC19

Given that the MR analyses used plasma protein levels in a non-infectious state as the exposures, we observationally assessed an association between plasma NPNT in a non-infectious state and the risk of COVID-19 severity outcomes in the BQC19 cohort. We defined the COVID-19 severity outcomes in accordance with those for the GWAS from COVID-19 Host Genetics Initiative (**Supplementary Information**).

We performed logistic regression analysis using one of the COVID-19 outcomes as the dependent variable and standardized plasma NPNT levels as the independent variable while adjusting for sex, age, and batch number. We did not adjust for clinical risk factors such as smoking and socioeconomic status.

### 4. Follow-up analyses for the putatively causal protein (NPNT)

#### 4.1. Colocalization of NPNT’s cis-pQTL with eQTL, and sQTL

##### sQTL and eQTL GWAS

We used sQTL and eQTL GWASs derived from the lung and those of thyroid and arteries (i.e., the top three *NPNT*-expressing tissues) in the European-ancestry individuals from the GTEx Portal V8 dataset^46^ (https://gtexportal.org/).

##### Colocalization analyses

To evaluate whether cis-pQTL is more affected by sQTL, rather than total expression, we also carried out colocalization of the cis-pQTL with eQTLs and sQTLs of NPNT using the GTEx V8 dataset. Colocalization analyses were performed in the 1 Mb region around the cis-pQTL for NPNT (rs34712979) using “coloc.abf()” function from the coloc v5.1.0^79^ with default priors of p1 = p2 = 10^−4^ and p12 = 10^−5^ (the definitions of p1, p2, and p12 can be found above). PP_shared_ > 0.8 was considered to indicate strong evidence of colocalization. To test the robustness of the results, we evaluated 27 different combinations of priors: p1 = c(10^−4^, 10^−5^, 10^−6^), p2 = c(10^−4^, 10^−5^, 10^−6^), p12 = c(10^−5^, 5 × 10^−6^, 10^−6^), as performed previously^18^. We also evaluated whether the lead cis-pQTL from the deCODE study (rs34712979), the FENLAND study (rs34712979), and the AGES Reykjavik study (rs78213340) were associated with the same splice pattern in the lung using GTEx. For eQTL and sQTL, we considered associations with *P* < 1 × 10^−7^ statistically significant, as did the deCODE study^26^.

#### 4.2. Single-cell RNA-sequencing data of SARS-CoV-2-infected lungs

To understand the *NPNT* expression pattern in the lungs of SARS-CoV-2 infected individuals, we obtained single-cell transcriptomic data of SARS-CoV-2-infected lungs by Delorey *et al*^47^ from the Single Cell Portal of the Broad Institute (https://singlecell.broadinstitute.org/single_cell/) (Accession ID: SCP1052). The data contained 106,792 single cells from the lungs of 16 autopsy donors aged 30 to older than 89 years who died due to COVID-19.

We reanalyzed the data focusing on NPNT expression status. We analyzed the gene expression matrix and the associated metadata using R v4.1.2 and Seurat R package v4.1.1^83^ (https://satijalab.org/seurat/). For visualization, NPNT expression levels were represented on a log-transcript per 10 thousand + 1, i.e., log (TP10K+1) scale. To cluster the cells, we adopted the clustering annotation from the original study^47^. To test whether *NPNT* expression was enriched in a cell type, we calculated the proportion of *NPNT*-expression cells in this cell type. Subsequently, we permuted the cell type labels 1,000 times and obtained the frequency (permutation p-value) of the same cell type containing the same or a larger proportion of *NPNT*-expression cells. Additionally, we compared the *NPNT* expression level between a target cell type and the other cell types using Wilcoxon rank-sum test. This enrichment analysis was performed separately in SARS-CoV-2-infected and uninfected cells.

#### 4.3. Mediation analysis

We undertook mediation analysis to calculate the proportion of the effect of BMI on critically ill COVID-19 mediated by NPNT using network MR (**Figure 7**). We used the product of coefficient method to estimate the NPNT-mediated effect (i.e., the effect of BMI on critically ill COVID-19 that was accounted for by NPNT) and the same instrumental variables and outcome GWASs from step 1 and step 2 MR.

First, we estimated the effect of BMI on NPNT, then multiplied this by the effect of NPNT on critically ill COVID-19. Subsequently, the proportion of the total effect of BMI on COVID-19 mediated by NPNT was estimated by dividing the NPNT-mediated effect (*β*_NPNT-to-severity_) by the total effect (*β*_BMI-to-severity_), as described previously^49,84^. Additionally, we evaluated whether NPNT mediated the effect of body fat percentage on COVID-19 hospitalization and the effect of body fat mass on critically ill COVID-19 and COVID-19 hospitalization.

We used the product of coefficients method without adjusting for the exposure (BMI or body fat percentage) when estimating the effect of the mediator on the outcome (*β*_NPNT-to-severity_) to avoid weak instrument bias. This approach was also used in previous studies^48,49,50^. We did so because the above-mentioned exposure adjustment requires multivariable MR using NPNT and BMI (or body fat percentage; fat mass) as exposures; however, there are only two instrumental variables for NPNT (cis-pQTL), but hundreds of instrumental variables for BMI. Thus, when using multivariable MR, which includes instrumental variables from both exposures in the model, the association between plasma NPNT levels and instrumental variables would be substantially weakened (i.e., the large number of instrumental variables of BMI would decrease the strength of the association between plasma NPNT and the genetic variants). Nevertheless, in sensitivity analyses, we performed mediation analyses with adjustment for the exposure when estimating *β*_NPNT-to-severity_ (i.e., Sobel test) (**Supplementary Table 20**). For multivariable MR, we performed data harmonization using the “mv_harmonise_data()” function from TwoSampleMR v.0.5.6^72^, followed by multivariable MR causal estimation using the “mv_multiple()” function from MVMR v0.3 (https://github.com/WSpiller/MVMR)^51^. We calculated conditional F-statistics and heterogeneity Q-statistics using the “strength_mvmr()” and “pleiotropy_mvmr()” functions, respectively, again from MVMR v0.3. To the best of our knowledge, the sample dataset of the exposure did not overlap (one was the plasma NPNT levels from the deCODE study, which is an Icelandic cohort, and another was from the obesity-related traits from UK Biobank); thus, we set “gencov” to be zero when calculating these measures following the instruction by the package^85^, as performed previously^86^. We also quantified directional pleiotropy with the MR-Egger intercept test with the “mr_mvegger()” function from MendelianRandomization v0.6.0^85^ (https://github.com/cran/MendelianRandomization).

#### 4.4. Multivariable MR of body fat and fat-free mass

##### Body fat and fat-free mass GWAS

We obtained GWAS of body fat and fat-free mass from UK Biobank using the IEU OpenGWAS project (https://gwas.mrcieu.ac.uk/). Accession ID for each GWAS was “ukb-b-19393” and “ukb-b-13354”, respectively.

##### NPNT GWAS

For GWAS of plasma NPNT levels, we used the same GWAS from the deCODE study^26^ as the one used in Step 1 MR, which measured plasma NPNT levels in 35,559 individuals of European ancestry by using the SomaLogic SomaScan assay v4.

##### COVID-19 severity outcomes

We used the same GWASs of critically ill COVID-19 and COVID-19 hospitalization from the COVID-19 Host Genetics Initiatives data release 7 (https://www.covid19hg.org/).

##### Multivariable MR

Since body fat mass and fat-free mass are genetically correlated with each other (*r* = 0.64)^87^, we performed multivariable MR to estimate the independent effect of body fat and fat-free mass on plasma NPNT levels or COVID-19 severity outcomes. For instrumental variables, we identified genome-wide significant and independent SNPs for body fat and fat-free mass using the same criteria as in step 1 MR (i.e., *P* < 5 × 10^−8^ for significance and *r*^*2*^ < 0.001 with a clumping window of 10 Mb for independence).

SNPs in the MHC region were excluded. After data harmonization, we undertook multivariable MR with the inverse variance weighted method using a random-effects model. We used body fat and fat-free mass as the exposures and plasma NPNT levels as the outcome. Results with *P* < 0.025 (0.05/2; Bonferroni correction) were considered statistically significant. LD clumping was performed using PLINK v1.9^73^. We performed data harmonization using the “mv_harmonise_data()” function from TwoSampleMR v.0.5.6^72^, followed by multivariable MR causal estimation using the “mv_multiple()” function from MVMR v0.3 (https://github.com/WSpiller/MVMR)^51^. For sensitivity analyses, we first calculated genetic covariance matrix for exposures (i.e., body fat mass and fat-free mass) using the “phenocov_mvmr()” function, and then used “strength_mvmr()” and “pleiotropy_mvmr()” functions from MVMR v0.3^51^ to calculate conditional F-statistics and Q-statistics, respectively. To calculate a phenotypic correlation matrix used in the “phenocov_mvmr()” function, we used metaCCA v1.22.0 (https://github.com/acichonska/metaCCA)^88^, as previously performed by Vabistsevits *et al*^86^. Lastly, to perform multivariable MR-Egger analysis, we used the “mr_mvegger()” function from MendelianRandomization v0.6.0^85^ (https://github.com/cran/MendelianRandomization).

## Data Availability

・GWAS summary statistics for each trait are available as follows: 
BMI (https://portals.broadinstitute.org/collaboration/giant/),
Plasma proteome from the deCODE study (https://www.decode.com/summarydata/), 
COVID-19 outcomes (https://www.covid19hg.org/results/r7/), 
GTEx Portal V8 (https://gtexportal.org/home/datasets/).
Plasma proteome from BQC19 (https://www.mcgill.ca/genepi/mcg-covid-19-biobank) Access to the data of BQC19 can be obtained upon approval of requests via bqc19.ca.
・Cis-pQTLs of each study are available in the corresponding publications' supplementary materials26-28
・Body fat percentage, body fat mass, and fat-free mass GWASs are available at IEU OpenGWAS project with Accession ID of ukb-b-8909, ukb-b-19393, and ukb-b-13354, respectively (https://gwas.mrcieu.ac.uk/)
・Single-cell RNA-sequencing data of COVID-19 lung autopsy samples are available at the Single Cell Portal under the Accession ID of SCP1052 (https://singlecell.broadinstitute.org/single_cell/) 
・CADD-scores v.1.6 can be accessed at https://cadd.gs.washington.edu/score
・Genotype data from 1000G genomes project is available at https://www.internationalgenome.org/data
Code availability
We used R v4.1.2 (https://www.r-project.org/), TwoSampleMR v.0.5.6 (https://mrcieu.github.io/TwoSampleMR/), snappy v1.0 (https://gitlab.com/richards-lab/vince.forgetta/snappy), coloc v5.1.0 (https://chr1swallace.github.io/coloc/), FINEMAP R package v1.4, Seurat v4.0.6 (https://satijalab.org/seurat/), PLINK v1.9 (http://pngu.mgh.harvard.edu/purcell/plink/), and GCTA fastGWA v1.93.3 (https://yanglab.westlake.edu.cn/software/gcta/). Custom codes are available on GitHub (https://github.com/satoshi-yoshiji/TwostepMR_obesity_COVID/).

## Ethical approval

For summary-level data, all contributing cohorts obtained ethical approval from their intuitional ethics review boards. The contributing cohorts include: UK Biobank, GIANT consortium, deCODE study, FENLAND study, AGES Reykjavik study, INTERVAL study, COVID-19 Host Genetics Initiative, and BQC19. For individual-level data in BQC19, BQC19 received ethical approval from the Jewish General Hospital research ethics board (2020-2137) and the Centre Hospitalier de l’Université de Montréal institutional ethics board (MP-02-2020-8929, 19.389). All participants provided informed consent.

## Data availability

GWAS summary statistics for each trait are available as follows:

BMI (https://portals.broadinstitute.org/collaboration/giant/),

Plasma proteome from the deCODE study (https://www.decode.com/summarydata/),

COVID-19 outcomes (https://www.covid19hg.org/results/r7/),

GTEx Portal V8 (https://gtexportal.org/home/datasets/).

Plasma proteome from BQC19 (https://www.mcgill.ca/genepi/mcg-covid-19-biobank)

Access to the data of BQC19 can be obtained upon approval of requests via bqc19.ca.

- Cis-pQTLs of each study are available in the corresponding publications’ supplementary materials^26-28^
- Body fat percentage, body fat mass, and fat-free mass GWASs are available at IEU OpenGWAS project with Accession ID of ukb-b-8909, ukb-b-19393, and ukb-b-13354, respectively (https://gwas.mrcieu.ac.uk/)
- Single-cell RNA-sequencing data of COVID-19 lung autopsy samples are available at the Single Cell Portal under the Accession ID of SCP1052 (https://singlecell.broadinstitute.org/single_cell/)
- CADD-scores v.1.6 can be accessed at https://cadd.gs.washington.edu/score
- Genotype data from 1000G genomes project is available at https://www.internationalgenome.org/data

## Code availability

We used R v4.1.2 (https://www.r-project.org/), TwoSampleMR v.0.5.6 (https://mrcieu.github.io/TwoSampleMR/), snappy v1.0 (https://gitlab.com/richards-lab/vince.forgetta/snappy), coloc v5.1.0 (https://chr1swallace.github.io/coloc/), FINEMAP R package v1.4, Seurat v4.0.6 (https://satijalab.org/seurat/), PLINK v1.9 (http://pngu.mgh.harvard.edu/purcell/plink/), and GCTA fastGWA v1.93.3 (https://yanglab.westlake.edu.cn/software/gcta/). Custom codes are available on GitHub (https://github.com/satoshi-yoshiji/TwostepMR_obesity_COVID/).

## Acknowledgments

We thank the COVID-19 Host Genetics Initiative for providing the latest summary statistics for COVID-19 outcomes. We acknowledge Shidong Wang, Tala Khosroheidari, Lena Cuddeback, Will Schwarzmann, and DeAunne Denmark at SomaLogic, Inc. for constructive discussions. We acknowledge Biorender (biorender.com) for providing materials used to create the illustrative diagram. The Richards research group is supported by the Canadian Institutes of Health Research (CIHR: 365825, 409511, 100558, 169303), the McGill Interdisciplinary Initiative in Infection and Immunity (MI4), the Lady Davis Institute of the Jewish General Hospital, the Jewish General Hospital Foundation, the Canadian Foundation for Innovation, the NIH Foundation, Cancer Research UK, Genome Québec, the Public Health Agency of Canada, McGill University, Cancer Research UK [grant number C18281/A29019] and the Fonds de Recherche Québec Santé (FRQS). J.B.R. is supported by an FRQS Mérite Clinical Research Scholarship. The support from Calcul Québec and Compute Canada is acknowledged. TwinsUK is funded by the Welcome Trust, Medical Research Council, European Union, the National Institute for Health Research (NIHR)-funded BioResource, Clinical Research Facility and Biomedical Research Centre based at Guy’s and St Thomas’ NHS Foundation Trust in partnership with King’s College London. S.Y. is supported by the Japan Society for the Promotion of Science. T.L. is supported by a Vanier Canada Graduate Scholarship, an FRQS doctoral training fellowship, and a McGill University Faculty of Medicine Studentship. G.B.L. is supported by scholarships from the FRQS, the CIHR, and Québec’s ministry of health and social services. Y.C. is supported by an FRQS doctoral training fellowship and the Lady Davis Institute/TD Bank Studentship Award. M.H. is supported by grants from the SciLifeLab/Knut and Alice Wallenberg national COVID-19 research program (M.H.: KAW 2020.0182, KAW 2020.0241), the Swedish Heart-Lung Foundation (M.H.: 20210089, 20190639, 20190637), and the Swedish Society of Medicine (M.H.:SLS-938101). The funders had no role in study design, data collection and analysis, decision to publish or preparation of the manuscript.

## Author contributions

Conception and design: S.Y. and J.B.R. Methodology: S.Y., T.L., and J.B.R. Data analysis: S.Y., T.L., J.D.S.W., C.Y.S., and J.B.R. Visualization: S.Y. and T.L. Writing – original draft: S.Y. Writing – Review and editing: S.Y., G.B.L., T.L., J.D.S.W., C.Y.S., T.N., D.M., Y.C., K.L., M.H., Y.I., Z.A., S.L., N.D., C.D., M.V., C.T., X.X., M.B., F.S., L.L., H.M.M., M.A., J.A., V.M., N.J.T., H.Z., S.Z., V.F., Y.F., and J.B.R.

