## Supplementary Information for "Proteome-wide Mendelian randomization implicates nephronectin as an actionable mediator of the effect of obesity on COVID-19 severity"

### Sample processing in the BQC19 cohort

The samples underwent proteomic profiling using the SomaScan v4 assay. Up to 5,284 aptamers were measured for these samples. After removing any aptamers that represented non-human proteins or controls, we retained 4,907 aptamers for analysis, consistent with the deCODE study[26]. SomaLogic performed normalization and calibration steps to remove systematic biases, which are detailed in their technical note (https://www.mcgill.ca/genepi/files/genepi/bqc19_jgh_prt_tech_note_0.pdf). Blood samples were sent to SomaLogic at two-time points during the pandemic, providing two batches of proteomic measurements. For quality control, the protein levels were natural log-transformed and subsequently batch-corrected using the ComBat function implemented in the sva R package v3.44.0[81], which uses an empirical Bayesian framework to perform batch effect removal.

### Definition of SARS-CoV-2 infectious or non-infectious state in the BQC-19 cohort

We defined infectious and non-infectious states as follows (all date ranges are inclusive): infectious samples were defined as blood samples collected from individuals who tested positive for SARS-CoV-2 test from 7 days before and up to, and including 14 days after the first date of SARS-CoV-2-associated symptoms. Non-infectious samples were defined as those meeting either of the following three criteria: (1) samples were collected from individuals who tested negative for SARS-CoV-2; (2) samples were collected within 31 days after, but 90 days before the first date of symptoms from patients whose positive SARS-CoV-2 test was confirmed within 7 days from the first symptom onset; (3) samples were collected at least 15 days before or 31 days after the positive SARS-CoV-2 test from patients whose positive SARS-CoV-2 test was confirmed at least 7 days before or after the first symptom onset. Further details can be found at https://github.com/richardslab/BQC19_phenotypeQC/blob/main/src/COVID-19%20Omics.svg.

### Definition of the COVID-19 severity outcomes in the BQC-19 cohort

We defined the COVID-19 outcomes in accordance with those for the GWAS from COVID-19 Host Genetics Initiative. (I) Critically ill COVID-19: cases were defined as laboratory-confirmed SARS-CoV-2 infection by PCR or serology testing along with the requirement for respiratory support or death. (II) Hospitalization: cases were defined as those who were hospitalized due to COVID-19-related symptoms. Controls were defined as individuals who tested negative for SARS-CoV-2.
